# Light Enhanced Cognitive Behavioral Therapy (CBT_+_) for Insomnia and Fatigue During Chemotherapy for Breast Cancer: A Randomized Controlled Trial

**DOI:** 10.1101/2021.03.23.21254147

**Authors:** Helena R. Bean, Justine Diggens, Maria Ftanou, Marliese Alexander, Lesley Stafford, Bei Bei, Prudence A. Francis, Joshua F. Wiley

## Abstract

**Study Objectives:** Sleep problems are common during chemotherapy for breast cancer (BC). We evaluated whether combined brief cognitive behavioral and bright light therapy (CBT_+_) is superior to treatment as usual with relaxation audio (TAU_+_) for insomnia symptoms and sleep efficiency (primary outcomes).

**Methods:** We randomized women receiving intravenous chemotherapy, stratified by tumor stage and insomnia severity index (ISI), to 6-weeks CBT_+_ or TAU_+_. CBT_+_ included one in-person session, one telephone call, seven emails, and 20 minutes bright light each morning. TAU_+_ comprised usual treatment and two emails with relaxation audio tracks. Patient-reported outcomes were assessed at baseline, midpoint (week 3), post (week 6) and 3-month follow-up.

**Results:** Women (*N* = 101) were randomly assigned to CBT_+_ or TAU_+_. Insomnia symptoms declined significantly more from baseline to post with CBT_+_ versus TAU_+_ (−5.06 vs -1.93, *P* = .009; effect size [ES] = .69). At 3-month follow-up, both groups had improved insomnia symptoms but did not differ (ES = .18, *P* = .56). CBT_+_ had higher patient-reported sleep efficiency than TAU_+_ after the start of intervention (*P* = .05) and more improvement in fatigue (ES = .59, *P* = .013) and daytime sleep-related impairment (ES = .61, *P* = .009) from baseline to post.

**Conclusions:** CBT_+_ had a clinically significant impact on insomnia and fatigue with moderate effect sizes. Results support offering cognitive behavioral therapy for insomnia and bright light therapy during chemotherapy for breast cancer to help manage sleep and fatigue.

**Clinical trial information:** Registered with the Australian New Zealand Clinical Trials Registry (http://anzctr.org.au/), Registration Number: ACTRN12618001255279

**Statement of Significance:** Poor sleep is prevalent after cancer diagnosis, especially during chemotherapy. Although cognitive behavioral therapy for insomnia (CBT-I) has been shown to be effective, including in cancer survivors, chemotherapy is unique with significant, ongoing, exogenous factors disrupting sleep (e.g., pain, nausea, and steroids used to manage side effects from chemotherapy). We showed statistically and clinically significant improvements in insomnia and fatigue symptoms in the CBT-I plus bright light therapy (CBT_+_) treatment condition compared to the control in women receiving chemotherapy for stage 1–4 breast cancer. Waiting until cancer treatment completion may result in prolonging poor sleep and our findings suggest is not necessary. Future studies are needed to evaluate whether combined CBT_+_ is superior to CBT-I or light alone.

## Introduction

Sleep disturbance affects up to 80% of women with breast cancer (BC)^1-4^ and a large, prospective study found 43% of people during chemotherapy met criteria for insomnia syndrome^5^. Three broad domains contribute to sleep disturbance in BC: (1) treatments (e.g., surgery, radiotherapy, chemotherapy, hormonal therapy) and their side effects^6^, (2) psychological challenges of diagnosis and treatment (e.g., stress, depression, and anxiety)^2,7,8^; and (3) circadian rhythm disruption^9-12^, with chemotherapy as a potential cause^13,14^. Although sleep complaints are reported throughout the BC trajectory^1^, the period surrounding chemotherapy is notable for the development and exacerbation of insomnia symptoms^15^. Untreated, sleep disturbance and insomnia symptoms are associated with physical and psychological consequences including: depression, anxiety, cognitive decline, impaired daily functioning, productivity and quality of life, and increased physical morbidity^4,16-18^. Despite the prevalence and consequences of poor sleep, there are few trials of sleep interventions during chemotherapy^19-21^.

To improve sleep in women with BC, multiple contributing domains need to be targeted. Cognitive Behavioral Therapy for Insomnia (CBT-I), the first-line treatment for insomnia^22^ including insomnia comorbid with cancer^4^, is a multi-component intervention that targets psychological and behavioral factors. CBT-I comprises sleep restriction, stimulus control, sleep hygiene, cognitive restructuring and can include relaxation training. CBT-I can be effective even when factors outside of an individual’s control (e.g., chemotherapy) interfere with sleep^2^. Further, there are benefits to a non-pharmacological approach, given the known consequences of pharmacological sleep-aid use^23^ and the potential for interactions between sleep-aids and chemotherapy drugs^24^. Limitations of CBT-I include delivery demands and limited accessibility^25-27^. A few trials have tested brief delivery formats of CBT-I in cancer (internet and video-based programs) showing efficacy^25,28^. However, CBT-I does not directly target circadian rhythm disruptions. Bright light (BL) therapy is a low-cost treatment for circadian rhythm disruption^29-31^. Three small studies showed that BL can prevent worsening fatigue and circadian rhythm deterioration during chemotherapy^32-34^; however, the impact of BL for sleep, during chemotherapy remains unclear.

To address these gaps, we developed a combined brief CBT-I and BL (CBT_+_) intervention that targets multiple mechanisms of poor sleep and conducted a randomized, controlled trial of CBT_+_ versus treatment as usual plus relaxation audios (TAU_+_) in women receiving chemotherapy for BC. Women with metastatic BC often have been excluded from previous sleep trials^19,21^ so that even less is known about the efficacy of sleep interventions in this high-risk group. We purposefully included metastatic BC to aid generalizability. The primary aim was to test the efficacy of CBT_+_ compared to TAU_+_ for the dual primary outcomes of patient reported insomnia symptoms and sleep efficiency (time asleep divided by time spent in bed for sleep). Secondary aims were to compare the effects of CBT_+_ versus TAU_+_ on sleep parameters from sleep diary and accelerometry and on patient reported outcomes (PROs) of fatigue, sleep-related impairment, sleep disturbance, and psychological symptoms. We hypothesized that women randomized to CBT_+_ (versus TAU_+_) would show greater improvements on the above outcomes.

## Methods

### Study Design

Detailed methods are in the published trial protocol^35^.Reporting follows CONSORT guidelines for social and psychological interventions and PROs. The Peter MacCallum Cancer Centre (PMCC) ethics committee (#17/159) approved the trial in accordance with the Declaration of Helsinki. This was a randomized, two-group, parallel, non-blinded, controlled, single-center, superiority trial, conducted at PMCC. After providing written informed consent and screening, eligible participants were randomized to CBT_+_ or TAU_+_ and commenced the 6-week intervention. PROs were assessed at baseline prior to the intervention, intervention mid-point (3 weeks), post-intervention (6 weeks) and 3-months post-intervention completion follow-up. Objective and self-report sleep parameters were measured continuously for the 6-week intervention period commencing the night after the first intervention session.

### Study participants

Eligible women were diagnosed with any stage of BC; aged ≥18 years; receiving intravenous chemotherapy at study entry; English speaking; able to access emails; able and willing to wear BL glasses. Exclusion criteria were male; receiving only neoadjuvant chemotherapy; and diagnosed with a severe psychiatric or substance use disorder via the MINI^36^ interview. History of migraines and very advanced (early), delayed (late), or highly variable sleep timing, or non-24 sleep and wake pattern, based on the Duke structured sleep interview^37^, were exclusions.

### Interventions

All intervention components were delivered by a provisional psychologist trained in CBT-I at Monash University Healthy Sleep Clinic. Sessions were recorded and reviewed by a senior CBT-I clinician.

CBT_+_ comprised one 60-minute face-to-face session at the PMCC Chemotherapy Day Unit; one 20-minute telephone call at study mid-point and seven, weekly CBT-I content emails. Light boxes and glasses produce comparable effects^38^. Participants wore Luminette® light glasses for 20 minutes (per manufacturer recommendation) each morning at the brightest setting (see Supplementary methods). Women reported the duration and timing of light in daily diaries. Reported light use ≥20min and within -1 to _+_4 hours of habitual rise time was considered adherent. CBT_+_ content included: general information and skills for better sleep (e.g., sleep hygiene, relaxation exercises, cognitive and behavioural strategies for dealing with night-time worries); fostering healthy attitudes and expectations about sleep following cancer diagnosis and during treatment; managing sleep challenges specific to cancer patients (e.g., physical discomfort, pain, daytime consequences of poor sleep); and identifying and managing symptoms of insomnia (e.g., stimulus control, bed restriction).

TAU_+_ was presented as an “audio relaxation group”. TAU_+_ comprised two emails with relaxation audio tracks developed by the Australian Cancer Council to assist in coping with cancer during the 6-week intervention period and a mid-point telephone call. Content included general relaxation strategies to be used at any time with no sleep-specific information. After the final follow-up, TAU_+_ participants received all CBT_+_ emails.

### Primary Outcomes

#### Insomnia Symptoms

The first primary outcome was change from baseline through mid-point to post intervention on the Insomnia Severity Index (ISI)^39^, widely used and validated, comprising 7 patient-reported items (e.g., “difficulty falling asleep”), rated from 0 (none) to 4 (very severe), yielding a total score from 0-28. Scores of 0-7 indicate no clinical insomnia, 8-14 subthreshold insomnia, 15-21 clinical insomnia, and 22-28 severe clinical insomnia^39^.

#### Sleep Efficiency

Our second primary outcome was Sleep Efficiency (SE_diary_), the ratio of self-reported total sleep time to total time spent in bed for sleep, from the consensus sleep diary^40^.

### Secondary Outcomes

#### Sleep Parameters

Self-reported sleep diaries and the wrist-worn accelerometer (ActiGraph model wGT3X-BT) measured sleep onset latency (SOL), time spent awake after sleep onset (WASO) and total sleep time (TST), which has shown minimal average bias compared to polysomnography for TST, SE, and WASO in multiple studies^41,42^. Accelerometry data scoring followed a standard protocol, integrating estimates from an algorithm^43^, ambient light, and sleep diaries.

#### Patient-Reported Outcomes Measurement System (PROMIS) scales

validated 8-item short-form PROMIS scales assessed sleep related impairment, sleep disturbance^44^, fatigue^45^, depression and anxiety symptoms^46^.

### Random Assignment

REDCap (Research Electronic Data Capture) was used by a study member (BB) not involved in recruitment, to generate a 1:1 randomization scheme in advance, stratified by ISI scores (≤7, ≥8) and cancer stage (≤2, ≥3). Random seeds were generated, and variable block sizes used to assure concealment.

### Statistical Analyses

Power analyses based on t-tests showed that 35 women completing each group would provide >80% power at α=.05 (two-sided) to detect a standardized mean difference of 0.70, a clinically meaningful difference.

Descriptive statistics are frequencies and percentages for discrete variables and means and standard deviations for continuous variables. Baseline group differences were tested using t-tests or chi-square tests to verify successful randomization. Due to non-normality, sleep efficiency was transformed as: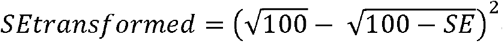 and SOL and WASO were square root transformed, which normalized their distributions. SOL and WASO were winsorized at the 0.5% percentiles address outliers.

Analyses followed a pre-specified plan^35^ and were intention-to-treat, conducted in R (v3.6) and MPlus (v8) with MplusAutomation^47^ on transformed variables. Tests were two-sided. Graphs are on the original scale. Analyses were piecewise latent growth models with an intercept and two linear slopes capturing (1) change from baseline through mid-point to post-treatment (primary hypothesis) and (2) maintenance from post-treatment to follow-up. Sleep diary and actigraphy analyses were mixed-effects models with a random intercept and linear slope of days since start of intervention. Analyses allowed freely estimated means, variances, and covariances of random intercepts and slopes and assumed a homogenous, independent residual variance. A constrained longitudinal analysis approach was taken^48,49^ and stratification factors were included as covariates^50,51^. Effect sizes (ES) between assigned groups were adjusted, standardized mean differences, standardized by the combined residual and intercept variance. Missing data were addressed using maximum likelihood^52^. Exploratory analyses examined results (1) in the subgroup with ISI≥8 at screening and (2) in sleep parameters by including weekday-weekend status and dexamethasone use as covariates.

## Results

### Patient Characteristics and Adherence

Between July 2018 and November 2019, 101 women receiving intravenous chemotherapy for breast cancer were randomized a median of 16 weeks post self-reported diagnosis (Figure 1). At randomization, 51.5% were stage ≤2 and 48.5% stage ≥3; 73.3% had subthreshold or more insomnia (ISI ≥8) and 26.7% had no clinical insomnia (ISI ≤7). There were no significant groups differences in sociodemographics, cancer treatments, or PROs at baseline (Table 1) nor in dropout across the study (Figure 1; additional results in Supplement). Self-reported credibility and expected benefit of the two interventions did not differ at baseline (see Supplement).

**Figure 1.**
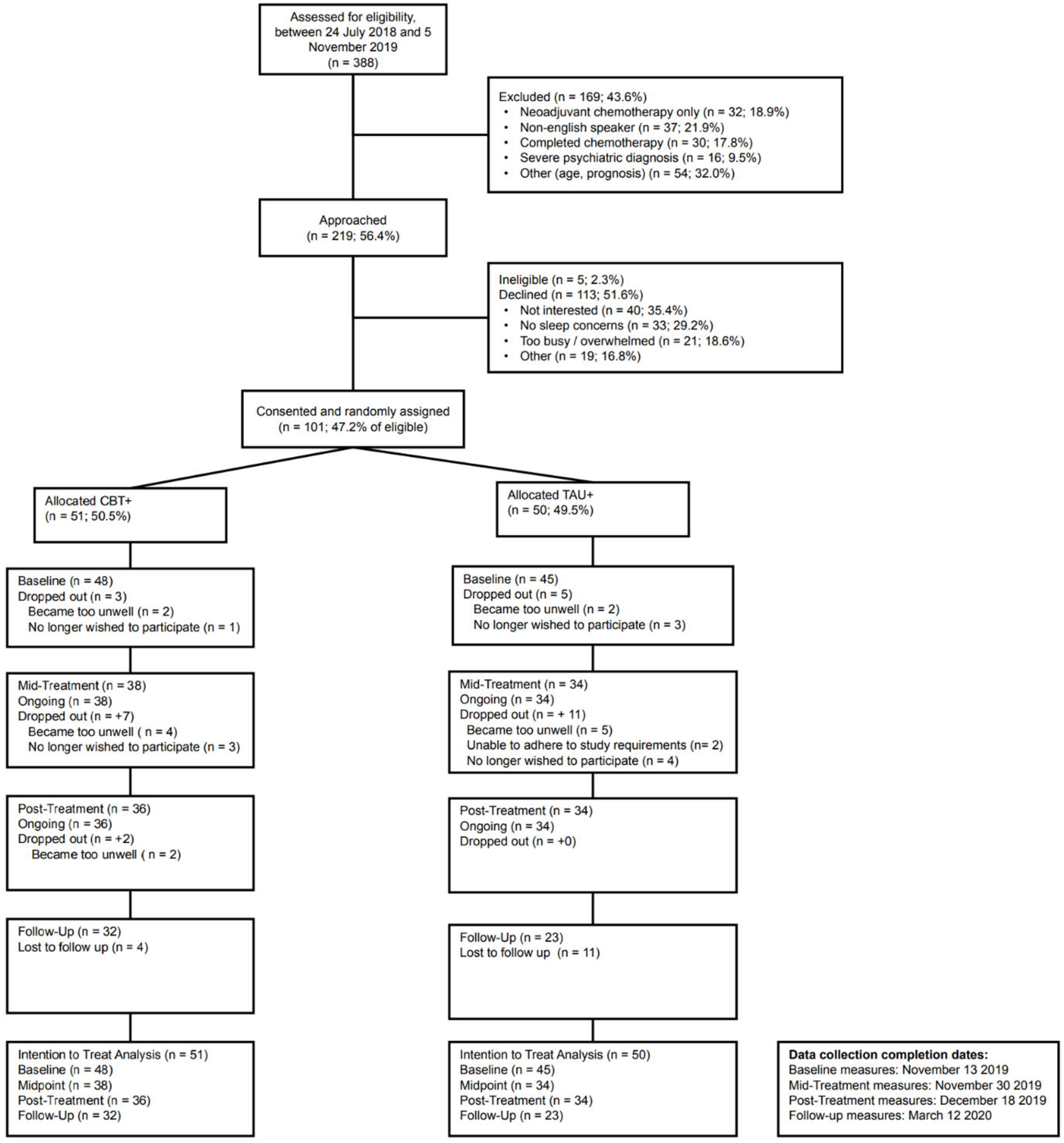
CONSORT diagram and recruitment flow chart.

**Table 1.** Baseline Sample Characteristics by Group

|  | CBT+ (n = 47) | TAU+ (n = 45) | P |
| --- | --- | --- | --- |
| Age (range: 27- 69 years), *M* (*SD*) | 48.52 (9.51) | 51.19 (12.02) | .24 |
| Married/living as married, *N* (%) | 33 (70.2) | 26 (57.8) | .21 |
| Children under 18, living at home, *N* (%) | 18 (38.3) | 10 (22.2) | .094 |
| Race/Ethnicity, *N* (%) |  |  | .14 |
| White/European | 41 (87.2) | 40 (88.9) |  |
| Asian | 2 (4.3) | 4 (8.8) |  |
| Other | 4 (8.5%) | 1 (2.2) |  |
| Income, *N* (%) |  |  | .12 |
| < AU\$50 000 | 7 (16.3) | 10 (24.4) | |
| AU\$50000-\$100000 | 15 (34.9) | 20 (48.8) | |
| > AU\$100000 | 21 (48.8) | 11 (26.8) | |
| Education, *N* (%) |  |  | .51 |
| < Bachelor | 14 (29.8) | 18 (40.0) |  |
| Bachelor | 17 (36.2) | 12 (26.7) |  |
| > Bachelor | 16 (34.0) | 15 (33.3) |  |
| Employment, *N* (%) |  |  | .62 |
| Employed | 32 (68.1) | 29 (64.4) |  |
| Retired | 6 (12.8) | 9 (20.0) |  |
| Not employed | 9 (19.1) | 7 (15.6) |  |
| Menopause, *N* (%) |  |  | .34 |
| Perimenopausal | 11 (26.2) | 10 (22.7) |  |
| Premenopausal | 20 (47.6) | 16 (36.4) |  |
| Postmenopausal | 11 (26.2) | 18 (40.9) |  |
| Cancer stage, *N* (%) |  |  | .68 |
| 1 | 5 (11.1) | 9 (20.5) |  |
| 2 | 16 (35.6) | 14 (31.8) |  |
| 3 | 12 (26.7) | 10 (22.7) |  |
| 4 | 12 (26.7) | 11 (25.0) |  |
| BC Surgery Prior to Baseline (Yes), *N* (%) | 33 (73.3) | 30 (68.2) | .59 |
| Radiotherapy, *N* (%) |  |  | .46 |
| No, Not Planned | 6 (13.6) | 12 (27.3) |  |
| No, Planned | 26 (59.1) | 21 (47.7) |  |
| No, Unknown to Patient if Planned | 7 (15.9) | 6 (13.6) |  |
| Yes Current | 5 (11.4) | 5 (11.4) |  |
| Hormonal Therapy, *N* (%) |  |  | .84 |
| No, Not Planned | 11 (25.6) | 13 (30.2) |  |
| No, Planned | 12 (27.9) | 11 (25.6) |  |
| No, Unknown to Patient if Planned | 14 (32.6) | 11 (25.6) |  |
| Yes Current | 6 (14.0) | 8 (18.6) |  |
| Mental Health Treatment since Diagnosis |  |  |  |
| Psychological (Yes), *N* (%) | 17 (38.6) | 13 (29.5) | .37 |
| Group or Other Support Resource (Yes), *N* (%) | 16 (35.6) | 9 (20.9) | .13 |
| Medication (Yes), *N* (%) | 9 (20.5) | 9 (20.9) | .96 |
| Sleep Treatment since Diagnosis |  |  |  |
| Psychological (Yes), *N* (%) | 5 (11.4) | 3 (6.8) | .46 |
| Over the Counter / Herbal Aids (Yes), *N* (%) | 10 (22.2) | 8 (18.2) | .64 |
| Prescribed Medication (Yes), *N* (%) | 17 (37.8) | 16 (36.4) | .89 |
| Insomnia Symptoms, *M* (*SD*) | 13.17 (5.61) | 12.86 (5.69) | .80 |
| Sleep-Related Impairment, *M* (*SD*) | 56.72 (7.93) | 56.86 (7.71) | .93 |
| Sleep Disturbance, *M* (*SD*) | 55.27 (9.36) | 54.74 (8.12) | .77 |
| Fatigue Symptoms, *M* (*SD*) | 57.37 (7.51) | 59.09 (7.77) | .29 |
| Depression Symptoms, *M* (*SD*) | 50.09 (8.67) | 51.89 (9.92) | .36 |
| Anxiety Symptoms, *M (SD)* | 52.83 (9.10) | 51.84 (10.14) | .63 |

*Note.* P-values come from independent sample t-tests for continuous variables (age) and chi-square tests for categorical variables. ES = effect sizes, which are Cohen's d for continuous variables and Cramer's V for categorical variables. Dx = breast cancer diagnosis; OTC = over the counter; Tx = treatment; N = 93 for baseline, however due to small amounts of missing data (e.g., declining to answer or not knowing the answer) sample size varies slightly by variable. Mental health treatment comprised any psychological treatment, support groups, or medication for mental health. Sleep treatment comprised any psychological, prescribed medication, or over the counter or herbal supplements for sleep. CBT+ = cognitive behavioral therapy + bright light therapy. TAU+ = treatment as usual + relaxation audios.

CBT_+_ and TAU_+_ had comparable intervention email open rates (84% vs 87%). On average, self-reported intervention strategy use was 3-4 times per week in both groups. BL adherence was 4.18 mornings per week counting missed daily reports as non-adherence or 6.28 mornings per week excluding missed reports. There were no reported adverse events deemed to be a direct result of trial participation.

### Primary Outcomes

All PROs had excellent reliability with Cronbach’s α ≥0.87 across all timepoints. Insomnia symptoms declined significantly in both groups from baseline to post-intervention; CBT_+_ declined -5.06 versus -1.93 in TAU_+_ (3.13 points more, *P* = .009; see Table 2). Figure 2 Panel A presents adjusted means, effect sizes, and group differences. To evaluate heterogeneity in treatment response, percent change from baseline to post-intervention in insomnia symptoms for each participant was calculated (Figure 3) showing that more women improved in CBT_+_ vs TAU_+_ (89% vs 68%), more women showed at least a 50% symptom reduction in CBT_+_ vs TAU_+_ (49% vs 15%), and fewer women had worsened insomnia symptoms in CBT_+_ vs TAU_+_ (9% vs 26%). From post-intervention to 3-month follow-up, there was no change in the CBT_+_ group, but the TAU_+_ group showed a further 2.52-point reduction (*P* = .023; Table 2). There were no group differences at 3-month follow-up (Figure 2 Panel A).

**Table 2.**
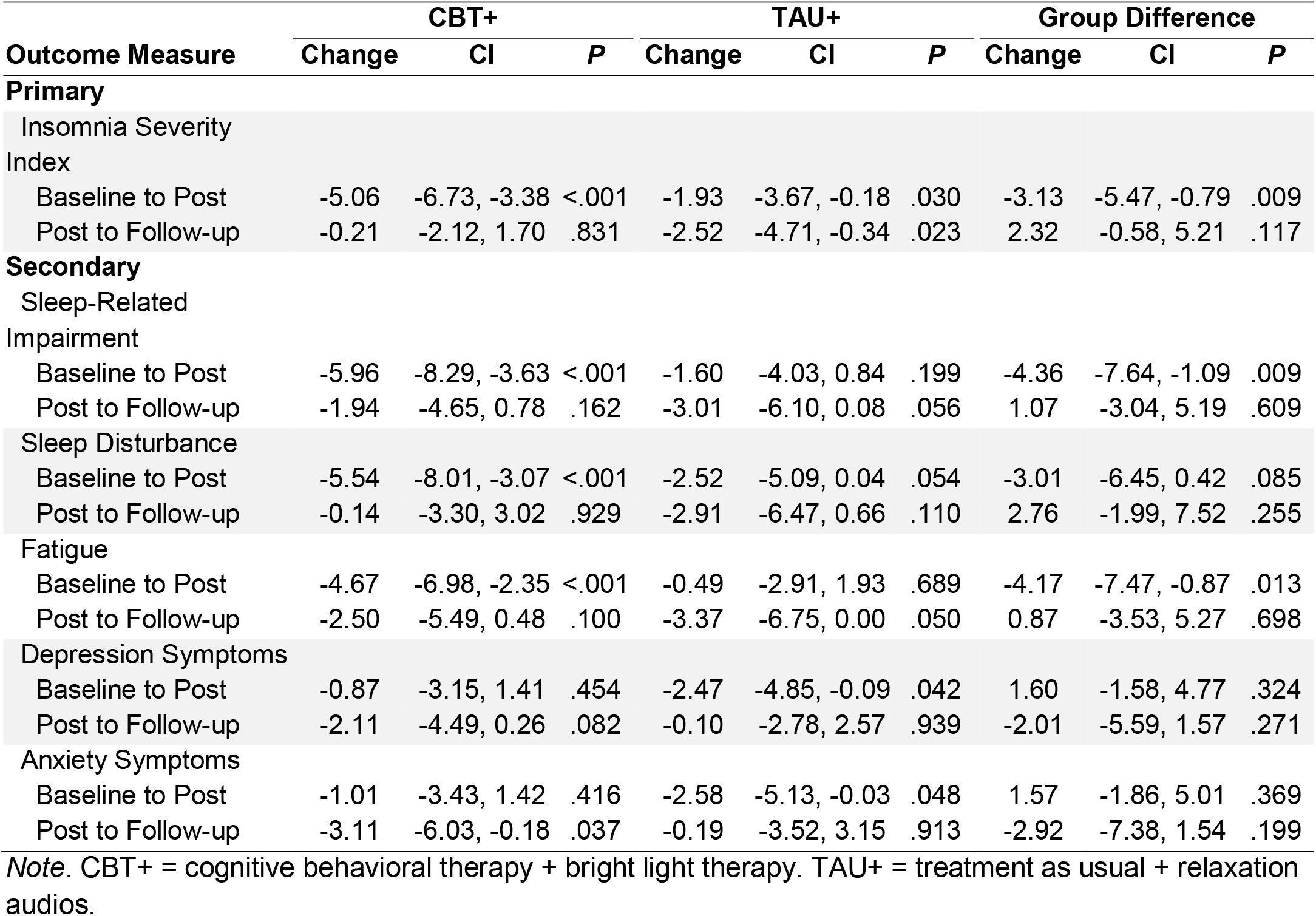
Changes by Treatment Group

| Outcome Measure | CBT+ |  |  | TAU+ |  |  | Group Difference |  |  |
| --- | --- | --- | --- | --- | --- | --- | --- | --- | --- |
|  | Change | CI | P | Change | CI | P | Change | CI | P |
| **Primary** |  |  |  |  |  |  |  |  |  |
| Insomnia Severity Index |  |  |  |  |  |  |  |  |  |
| Baseline to Post | -5.06 | -6.73, -3.38 | <.001 | -1.93 | -3.67, -0.18 | .030 | -3.13 | -5.47, -0.79 | .009 |
| Post to Follow-up | -0.21 | -2.12, 1.70 | .831 | -2.52 | -4.71, -0.34 | .023 | 2.32 | -0.58, 5.21 | .117 |
| **Secondary** |  |  |  |  |  |  |  |  |  |
| Sleep-Related Impairment |  |  |  |  |  |  |  |  |  |
| Baseline to Post | -5.96 | -8.29, -3.63 | <.001 | -1.60 | -4.03, 0.84 | .199 | -4.36 | -7.64, -1.09 | .009 |
| Post to Follow-up | -1.94 | -4.65, 0.78 | .162 | -3.01 | -6.10, 0.08 | .056 | 1.07 | -3.04, 5.19 | .609 |
| Sleep Disturbance |  |  |  |  |  |  |  |  |  |
| Baseline to Post | -5.54 | -8.01, -3.07 | <.001 | -2.52 | -5.09, 0.04 | .054 | -3.01 | -6.45, 0.42 | .085 |
| Post to Follow-up | -0.14 | -3.30, 3.02 | .929 | -2.91 | -6.47, 0.66 | .110 | 2.76 | -1.99, 7.52 | .255 |
| Fatigue |  |  |  |  |  |  |  |  |  |
| Baseline to Post | -4.67 | -6.98, -2.35 | <.001 | -0.49 | -2.91, 1.93 | .689 | -4.17 | -7.47, -0.87 | .013 |
| Post to Follow-up | -2.50 | -5.49, 0.48 | .100 | -3.37 | -6.75, 0.00 | .050 | 0.87 | -3.53, 5.27 | .698 |
| Depression Symptoms |  |  |  |  |  |  |  |  |  |
| Baseline to Post | -0.87 | -3.15, 1.41 | .454 | -2.47 | -4.85, -0.09 | .042 | 1.60 | -1.58, 4.77 | .324 |
| Post to Follow-up | -2.11 | -4.49, 0.26 | .082 | -0.10 | -2.78, 2.57 | .939 | -2.01 | -5.59, 1.57 | .271 |
| Anxiety Symptoms |  |  |  |  |  |  |  |  |  |
| Baseline to Post | -1.01 | -3.43, 1.42 | .416 | -2.58 | -5.13, -0.03 | .048 | 1.57 | -1.86, 5.01 | .369 |
| Post to Follow-up | -3.11 | -6.03, -0.18 | .037 | -0.19 | -3.52, 3.15 | .913 | -2.92 | -7.38, 1.54 | .199 |

*Note.* CBT+ = cognitive behavioral therapy + bright light therapy. TAU+ = treatment as usual + relaxation audios.

**Figure 2.**
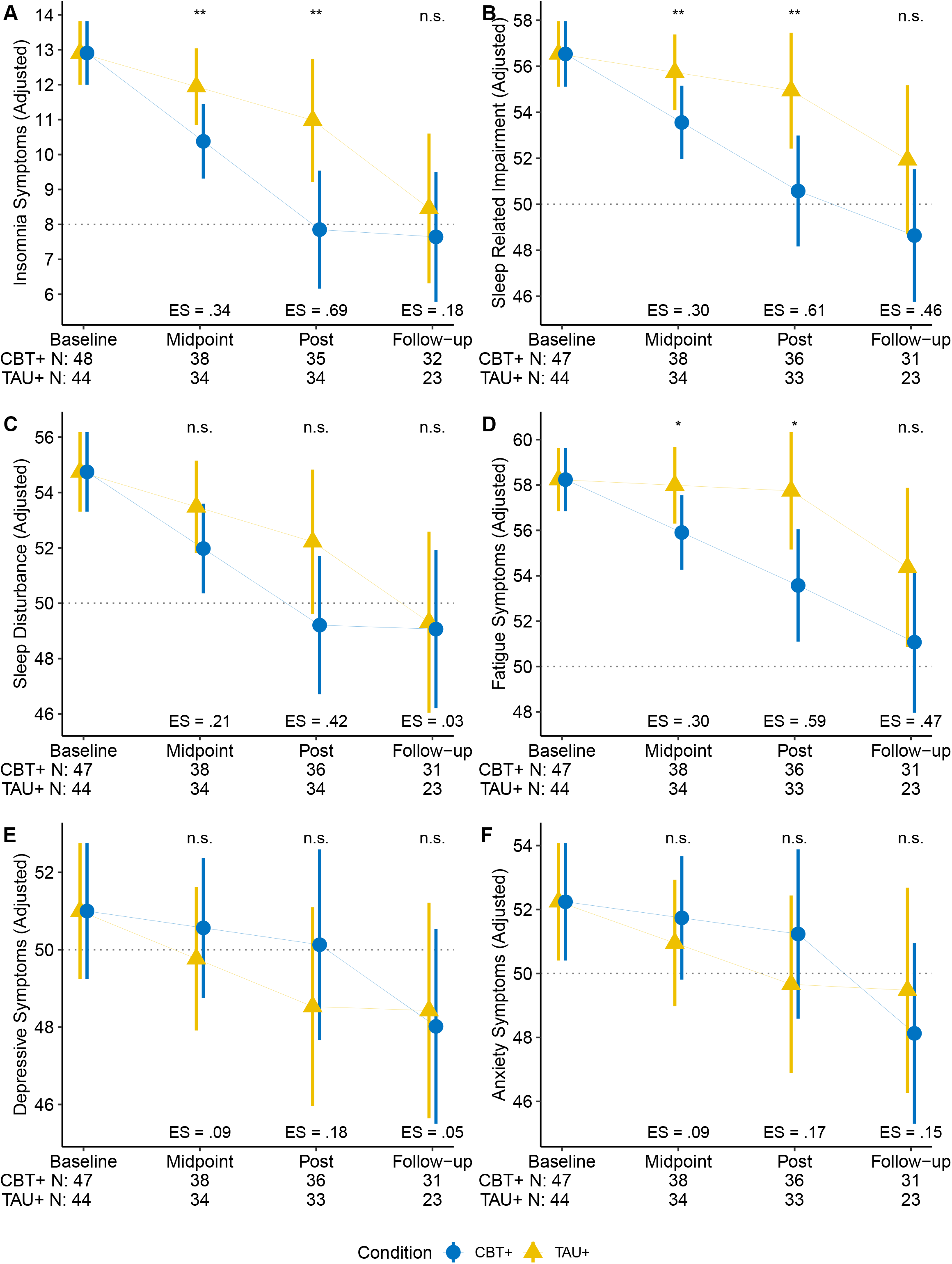
Changes in mean patient reported outcome measures at baseline, mid-point, post-intervention and 3-month follow up by group. Insomnia Symptoms (Panel A) is the primary outcome, all others (B - F) are secondary outcomes. Values are adjusted means and 95% confidence intervals from the primary analyses. Dashed grey line indicates population average values, where available, or minimal symptoms (insomnia symptoms only). The number of non-missing scores available in each group at each assessment are at the bottom. Baseline was completed just prior to week 0 (intervention start), midpoint at week 3, post at week 6, and follow-up 3-months after the completion of the intervention. CBT_+_ = cognitive behavioral therapy _+_ bright light therapy. TAU_+_ = treatment as usual _+_ relaxation audios. ES = effect size; *n*.*s*. = not significant; * *p* < .05; ** *p* < .01

**Figure 3.**
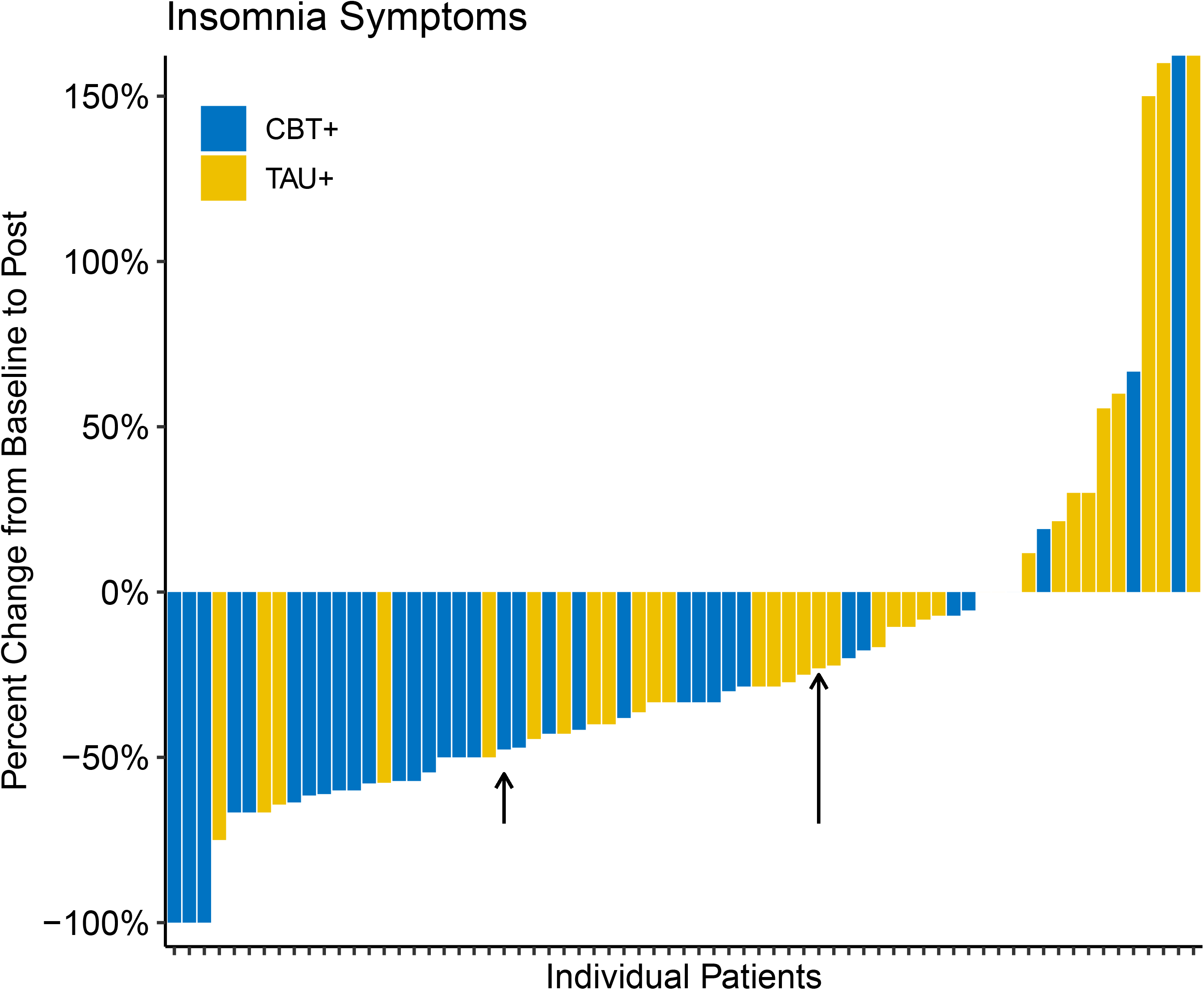
Individual participant change scores for insomnia symptoms from baseline (just prior to week 0, the intervention start) to post intervention (week 6) assessment points. CBT_+_ = cognitive behavioral therapy _+_ bright light therapy. TAU_+_ = treatment as usual _+_ relaxation audios. Arrows indicate the median change for CBT_+_ and TAU_+_.

Our second primary outcome, SE_diary_, was significantly higher in the CBT_+_ vs TAU_+_ group after the start of the intervention (*P* = .05; Online Table 3; Figure 4 Panel A). Both groups increased significantly over time (both *P* < .05) and this change did not differ between groups (*P* = .33). Although groups did not differ at the end of the intervention, both had average SE_diary_ > 85%, indicative of good sleep efficiency.

**Figure 4.**
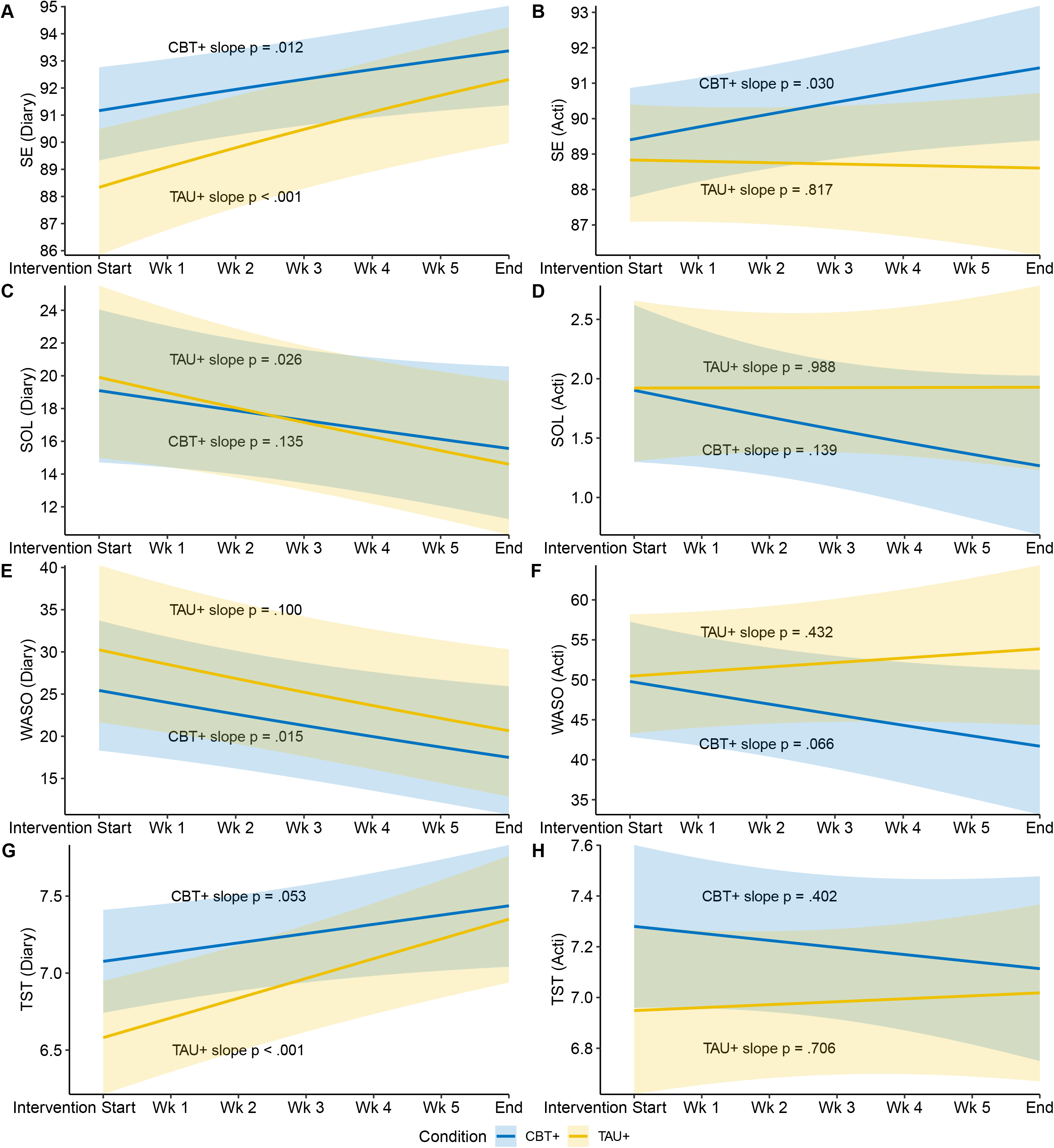
Changes in subjective and objective sleep from start to end of intervention period by group. SE_diary_ (Panel A) was the primary outcome. All others (B – H) were secondary outcomes. Sleep diary and actigraphy assessments began the first night after the initial face-to-face intervention session. Graphs are estimated means over time with 95% confidence intervals shown in the shaded regions. Diary = sleep measures assessed by sleep diary. Acti = sleep measures assessed by wrist-worn actigraphy. SE = sleep efficiency, in percentages. WASO = wake after sleep onset, in minutes. SOL = sleep onset latency, in minutes. TST = total sleep time, in hours. CBT_+_ = cognitive behavioral therapy _+_ bright light therapy. TAU_+_ = treatment as usual _+_ relaxation audios.

### Secondary Outcomes

#### Sleep and Psychosocial PRO

Changes over time and group differences for secondary PROs are in Table 2, with adjusted means, effect sizes and group differences in Figure 2 Panels B–F. When compared with TAU_+_, those assigned CBT_+_ improved 4.17 points more in fatigue, which was clinically meaningful and significant at 6-week post intervention time point (ES = .59, *P* = .013). Improvements in CBT_+_ over TAU_+_ also emerged for daytime sleep-related impairment (*P* = .009) with a trend for greater improvement in sleep disturbance (*P* = .085). Depression and anxiety symptoms did not differ between groups (both *P* > .30).

#### Sleep diary outcomes

SOL_diary_ and WASO_diary_ did not differ after the first intervention session and did not differ in how they changed over time (Online Table 3; Figure 4 Panels C & E). The trend was for both to decrease across the intervention. Compared to TAU_+_, the CBT_+_ group reported a trend towards 30min longer sleep duration (*P* = .051) after the start of the intervention and no differences in change over time (Online Table 3; Figure 4 Panel G).

#### Actigraphy outcomes

SE_acti_ did not differ between groups after the first intervention session, but there was a trend for different change trajectories over time (*P* = .080) with SE_acti_ increasing across the intervention for the CBT_+_ (*P* = .030) but not the TAU_+_ group (*P* = .817; see Online Table 3; Figure 4 Panel B). A similar pattern was observed for WASO_acti_, (Online Table 3 and Figure 4 Panel F). There were no differences after the first intervention session nor in the change over time for SOL_acti_ nor TST_acti_ (Online Table 3; Figure 4 Panels D & H).

### Exploratory Analyses

Exploratory analyses were conducted to examine whether treatment was more effective in subgroup of women with higher initial symptoms (*n* = 67 reported at least subthreshold insomnia with ISI ≥8 at screening). See detailed results in the Supplement (Online Tables 4 & 5; Online Figures 5 & 6). Improvements in insomnia symptoms remained significant and effect sizes were within ±.05 of results on overall sample, indicating minimal differences in benefits. Results for SE_diary_ also were comparable although in this subgroup, differences after the first intervention session were not significant. Improvements in fatigue in CBT_+_ vs TAU_+_ at post-intervention were larger (ES = .76, *P* = .005). Results for secondary outcomes were comparable.

Additional exploratory analyses on the sleep diary and actigraphy outcomes included two other sleep impacting factors as covariates: (1) weekend versus weekday sleep patterns, and (2) use of dexamethasone as part of chemotherapy regimen. Results were comparable to the overall sample with larger differences between CBT_+_ and TAU_+_ on SE_diary_ (*P* = .009) and TST_diary_ (*P* = .010) after the first intervention session in favor of CBT_+_. Rates of mental health and sleep treatment, including medication, receipt during the intervention did not differ between groups, nor were associated with change in insomnia symptoms. Detailed results are in the Supplement (Online Tables 6 & 7; Online Figure 7).

## Discussion

In women receiving chemotherapy for BC, this randomized trial examined the effects of CBT_+_ versus TAU_+_ on insomnia symptoms and sleep efficiency, as well as secondary outcomes of fatigue, other sleep parameters, and psychological symptoms. Women receiving CBT_+_ showed significantly greater improvements in the primary outcomes of insomnia symptoms and sleep efficiency, and secondary outcomes of sleep-related impairment and fatigue, supporting its efficacy as a sleep treatment in this population. Intervention effects on insomnia, fatigue and sleep-related impairment were clinically significant by 6 weeks (post-intervention), with women in the CBT_+_ group having mild insomnia symptoms that no longer differed from the population average, and fatigue symptoms improving beyond the 3-4 points considered a meaningful change, while the TAU_+_ group had no meaningful change in fatigue. At an individual level, all but three women in CBT_+_ experienced a decline in insomnia symptoms suggesting that CBT_+_ works, or at least does no harm, for nearly all women.

Exploratory analyses revealed that CBT_+_ is effective for women receiving chemotherapy for BC regardless of initial symptoms. Smaller improvements were observed in the TAU_+_ group for insomnia symptoms, sleep disturbance, and sleep efficiency, suggesting that TAU_+_ may provide some benefit, although we cannot distinguish effects of TAU_+_ versus natural declines in sleep problems over time.

Three months after ceasing intervention (including BL), CBT_+_ treatment benefits were maintained. Although outcomes were not significantly different between groups at follow-up, improvements in insomnia, sleep-related impairment, and fatigue via CBT_+_ over the 6-week intervention period may result in better functioning and quality of life during chemotherapy, a period when many other symptoms are distressing and difficult to alter.

Despite need and calls for greater recognition of sleep in cancer^53^, few trials target insomnia symptoms during chemotherapy. To our knowledge, no trial to date has combined brief CBT-I with BL to target the multiple causes of poor sleep during chemotherapy. A recent trial examined the effects of 21 days of bright light therapy only in this population and showed some benefit for sleep, depression and quality of life, however improvements in subjective sleep quality and fatigue were not statistically significant^54^. Comparatively, outcomes of the present trial may be indicative of the benefit in combining CBT-I and light therapy, or a longer intervention duration. In addition, approximately one quarter of participants in the CBT_+_ trial had metastatic BC, a group traditionally underrepresented in cancer sleep trials. Our outcomes and adherence data suggest that insomnia symptoms and fatigue can be treated during chemotherapy with a non-pharmacologic approach, despite the substantial physical and psychological impacts of active cancer treatment. CBT_+_ delivered using a combination of face-to-face (readily adaptable to telehealth delivery), telephone, and self-help materials is less burdensome on clinicians (about 1.5 hours) and patients than standard CBT-I (typically 6–10 hours). BL can be used at home with brief instruction and equipment mailed if needed, making CBT_+_ deliverable in cancer treatment clinics or at home in a scalable manner.

Strengths of this study include an intervention empirically grounded in the multiple determinants of insomnia symptoms and poor sleep during chemotherapy, random assignment, a manualized intervention, defined eligibility criterion, 6-weeks of subjective and objective sleep assessment, and equal representation of all stages of BC. This study also had limitations. To minimize burden, accelerometry and sleep diaries collection started after the first intervention session, so a true baseline was not available for these measures. Although accelerometry corresponds well with polysomnography for sleep duration, it tends to miss wake after sleep and likely underestimated sleep onset latency. Most participants were white, well-educated and had relatively high household income. Interventions were conducted by one interventionist at a single center. Future trials are needed with greater representation of race and ethnic groups and level of socioeconomic status and across multiple centers and interventionists to evaluate generalizability. About half of eligible women declined, primarily due to not being interested, no sleep concerns, or being overwhelmed. There was dropout after baseline, primarily from women reporting being too unwell, which may bias results. Targeting women with baseline symptoms or emphasizing the potential benefits of the trial may increase uptake. Although the trial was powered for its primary outcomes, larger trials would allow evaluating treatment efficacy in subgroups (e.g., chemotherapy cycle, cancer stage). Future trials are needed to evaluate whether CBT_+_ works in other cancer types and males. This first trial of CBT_+_ aimed to evaluate its efficacy, but future trials could investigate brief CBT-I and BL separately and together to identify any independent and synergistic effects.

In conclusion, this trial provides evidence that brief CBT_+_ can treat insomnia symptoms, sleep-related impairment, and fatigue for women with BC during chemotherapy. Chemotherapy can cause acute sleep disruption and fatigue adding to burden during a challenging time, so early intervention is crucial^4,19^.Our results support the feasibility and efficacy of intervening during chemotherapy and is one of the first to combine CBT-I and light therapy. Further research needs to consolidate findings, yet these results are an important step toward incorporating accessible, effective treatment for sleep into routine cancer care. Routine delivery of CBT_+_ may reduce the number of people with BC suffering from insomnia symptoms, fatigue, and associated impairment during and after chemotherapy.

## Supporting information

CONSORT Checklist

## Data Availability

Data will be shared upon reasonable request to the corresponding author.

## NOTES

All authors declare no conflict of interest. The funders had no role in the study design, data collection, analysis, interpretation, or presentation of results.

Data will be shared upon reasonable request to the corresponding author.

