## Supplementary material for "Light Enhanced Cognitive Behavioral Therapy (CBT_+_) for Insomnia and Fatigue During Chemotherapy for Breast Cancer: A Randomized Controlled Trial": Online Supplement

**Supplementary Methods – Light Therapy**

The light therapy glasses provided were Luminette® version 2. The light glasses were fixed at the brightest factory setting. Independent testing in our lab showed this provided white light, with a Correlated Colour Temperature (CCT) of 5,260 Kelvin. The peak spectral wavelength was at 466nm with an intensity of 1250 lux directed at each eye measured via spectrophotometry. The peak of this wavelength is very close to 480nm, the approximate peak sensitivity for melanopsin, a key mechanism of the effects of light on the circadian system and sleep. The melanopic lux of the light glasses was 813 lux, calculated using the International Commission on Illumination system (CIE) 2018.

**Supplementary Results – Dropout**

Participants lost to follow-up failed to respond to two phone calls and emails. At eligibility screening, there were no significant differences between dropouts and completers on stage (57.1% ≥ 3 in those who dropped out, 45.2% ≥ in completers; *P* = .28) or insomnia symptoms (82.1% ≥ 8 in those who dropped out, 69.9% ≥ 8 in completers; *P* = .21) or assigned group (42.9% CBT+ in those who dropped out, 53.4% in completers; *P* = .34). Of women who completed the baseline assessment, there were no difference in depression (*P* = .37) or anxiety (*P* = .32) symptoms between those who subsequently dropped out and those who completed, although in both cases, the direction of results was higher symptoms in those who dropped out. Overall, although none of the factors examined differed between women who completed or dropped out, the pattern was that women who dropped out were more likely to have more advanced stage, higher insomnia symptoms, worse mental health, and to be allocated to TAU+ rather than CBT+. A planned *a priori*, initial stage (≤ 2, ≥ 3) and insomnia symptoms (ISI ≤ 7, ≥ 8) were included as covariates in analyses, which should help to mitigate potential confounding.'

**Supplementary Results – Treatment Credibility and Expectancy**

Women's self-reports of expected benefit from the study interventions did not differ between groups at baseline, by which point women knew to which group they were assigned but had not yet begun the intervention (CBT+: M = 0.05, SD = 0.86; TAU+: M = -0.06, SD = 1.07; *P* = .606). Likewise, they did not differ in how credible and logical the interventions were (CBT+: M = 6.04, SD = 1.47; TAU+: M = 5.47, SD = 1.60; *P* = .082).

These results were based on the Credibility and Expectancy Questionnaire (Devilly & Borkovec, 2000; *Journal of Behavior Therapy and Experimental Psychiatry*). Three questions are about treatment credibility (e.g., "at this point, how logical does the program seem?") rated on a 9-point scale and three are about expectations for benefit (e.g., "By the end of the program, how much improvement in your sleep and daily functioning do you think will occur?"). Some expectancy items are rated on a 9-point Items and others on an 11-point scale, so the questionnaire authors recommend scaling the items about expectancy to a mean of zero and standard deviation of one, prior to average them to get an overall treatment expectancy rating.

**ONLINE ONLY Table 3.** Actigraphy and Sleep Diary Outcomes

| | **Sleep Diary B (95% CI)** | ***P*** | **Actigraphy B (95% CI)** | ***P*** |
|---|---|---|---|---|
| **Sleep Efficiency*** | | | | |
| *Intercept* | | | | |
| TAU+ | 43.36 (38.89, 47.83) | <.001 | 44.33 (41.04, 47.63) | <.001 |
| CBT+ | 49.39 (45.33, 53.44) | <.001 | 45.49 (42.29, 48.69) | <.001 |
| Group difference | 6.03 (-0.01, 12.07) | .050 | 1.15 (-3.44, 5.75) | .623 |
| *Slope* | | | | |
| TAU+ | 1.48 (0.74, 2.21) | <.001 | -0.08 (-0.71, 0.56) | .817 |
| CBT+ | 0.96 (0.21, 1.70) | .012 | 0.76 (0.07, 1.44) | .030 |
| Group difference | -0.52 (-1.57, 0.53) | .329 | 0.83 (-0.10, 1.77) | .080 |
| **Sleep Onset Latency*** | | | | |
| *Intercept* | | | | |
| TAU+ | 4.46 (3.87, 5.05) | <.001 | 1.39 (1.14, 1.63) | <.001 |
| CBT+ | 4.37 (3.84, 4.90) | <.001 | 1.38 (1.14, 1.62) | <.001 |
| Group difference | -0.09 (-0.89, 0.70) | .821 | -0.01 (-0.35, 0.34) | .969 |
| *Slope* | | | | |
| TAU+ | -0.11 (-0.20, -0.01) | .026 | 0.00 (-0.05, 0.05) | .988 |
| CBT+ | -0.07 (-0.16, 0.02) | .135 | -0.04 (-0.10, 0.01) | .139 |
| Group difference | 0.04 (-0.10, 0.17) | .594 | -0.04 (-0.12, 0.03) | .271 |
| **Wake after sleep onset*** | | | | |
| *Intercept* | | | | |
| TAU+ | 5.67 (4.79, 6.55) | <.001 | 7.10 (6.58, 7.63) | <.001 |
| CBT+ | 5.12 (4.33, 5.91) | <.001 | 7.06 (6.55, 7.57) | <.001 |
| Group difference | -0.55 (-1.73, 0.63) | .363 | -0.05 (-0.78, 0.69) | .900 |
| *Slope* | | | | |
| TAU+ | -0.10 (-0.23, 0.04) | .100 | 0.04 (-0.06, 0.14) | .432 |
| CBT+ | -0.16 (-0.28, -0.03) | .015 | -0.10 (-0.21, 0.01) | .066 |
| Group difference | -0.05 (-0.23, 0.13) | .570 | -0.14 (-0.28, 0.01) | .058 |
| **Total Sleep Time (hours)** | | | | |
| *Intercept* | | | | |
| TAU+ | 6.58 (6.21, 6.95) | <.001 | 6.95 (6.62, 7.28) | <.001 |
| CBT+ | 7.08 (6.74, 7.41) | <.001 | 7.28 (6.96, 7.60) | <.001 |
| Group difference | 0.50 (-0.00, 0.99) | .051 | 0.33 (-0.13, 0.79) | .158 |
| *Slope* | | | | |
| TAU+ | 0.13 (0.07, 0.19) | <.001 | 0.01 (-0.05, 0.07) | .706 |
| CBT+ | 0.06 (-0.00, 0.12) | .053 | -0.03 (-0.09, 0.04) | .402 |
| Group difference | -0.07 (-0.15, 0.02) | .119 | -0.04, (-0.13, 0.05) | .384 |

*Note*. *these measures were transformed, and results are on the transformed scales. CBT+ = cognitive behavioral therapy + bright light therapy. TAU+ = treatment as usual + relaxation audios.

### Exploratory Analyses – Elevated Screening Symptom Subgroup Analysis

An insomnia symptom threshold was not included in study eligibility criteria, allowing any otherwise eligible woman to participate, irrespective of her current sleep quality. Whilst the mean baseline ISI score of 12.91 indicated a moderate severity of insomnia symptoms in the sample, not all women reported significant insomnia symptoms. Analyses were conducted in the subset of women who reported baseline ISI scores of ≥ 8 to assess whether treatment effects differed. An ISI score of ≥ 8 has previously been determined as the optimal threshold for detecting clinically significant sleep difficulties in cancer populations with minimal false positive results [1,2]. The probability that patients with ISI scores ≥ 8 meet criteria for an insomnia diagnosis has been reported as 68% [1].

#### *Primary Outcomes*

As in the overall sample, insomnia symptoms declined significantly in both conditions from baseline to post intervention (both $P < .01$), and CBT+ declined 3.17 points more than TAU+ ($P = .025$; see Online Table 4). Online Figure 5 Panel A presents adjusted means, effect sizes, and group differences. Again as in the overall sample, from post-treatment to 3-month follow-up, there were no changes in the CBT+ condition, but women in TAU+ showed a further 3.49-point reductions ($P = .022$; Online Table 4), such that there were no group differences at 3-month follow-up (Online Figure 5 Panel A). Our second primary outcome was sleep efficiency, percent time in bed actually asleep, measured by daily sleep diaries ($SE_{diary}$) over the 6-week intervention period. Although the direction and magnitude were similar to the overall sample results, there were no significant difference in initial level or change over time in $SE_{diary}$ (Online Figure 6 and Online Table 5). Both groups increased significantly over time (both $P < .01$) and this change did not differ between groups ($P = .87$). As before, both groups had average $SE_{diary}$ > 85%, indicative of good sleep efficiency.

#### *Secondary Outcomes*

*Patient reported sleep and psychosocial outcomes*

Changes over time and group differences in these for secondary patient-reported outcomes in the initially high insomnia symptom subgroup are in Online Table 4, with adjusted means, effect sizes and group differences in Online Figure 5 Panels B – F.

CBT+ vs TAU+ had significantly greater improvements in fatigue (*P* = .005) with marginally significant improvements in sleep related impairment (*P* = .073). Sleep disturbance and depression and anxiety symptoms did not change significantly between groups (all *P* > .25). Compared to the overall sample, there were slightly stronger effects for fatigue and slightly weaker effects for sleep-related impairment and sleep disturbance.

*Sleep diary outcomes*

$SOL_{diary}$ and $WASO_{diary}$ did not differ after the first intervention session and did not differ in how they changed over time (Online Table 5; Online Figure 6 Panels C & E), although the trend was for both to decrease across the intervention (the decrease was significant in both cases for CBT+ but not TAU+). After the first intervention session, the CBT+ group reported about half an hour longer sleep duration than the TAU+ group as in the overall sample, although this was not significant in this smaller subsample (*P* = .120). There were no differences in change across the intervention (Online Table 5; Online Figure 6 Panel G).

*Actigraphy outcomes*

$SE_{acti}$ did not differ between groups after the first intervention session, but there was a marginally significant difference in change over time (*P* = .078) with $SE_{acti}$ increasing across the intervention for the CBT+ (*P* = .033) but not the TAU+ (*P* = .728), shown in Online Table 5 and Online Figure 6 Panel B. A similar pattern was observed for $WASO_{acti}$, (Online Table 5 and Online Figure 6 Panel F) and here the change over time did reach significance between groups (*P* = .017). There were no differences after the first intervention session nor in the change over time for $SOL_{acti}$ (Online Table 5 and Online Figure 6 Panel D). $TST_{acti}$ did not differ initially and neither group changed significantly over time, but the difference in change over time was significant (*P* = .019). The trend was for the CBT+ group to start slightly higher and decrease slightly whereas the TAU+ group started slightly lower and increased slightly and (Online Table 5 and Online Figure 6 Panel H).

**ONLINE ONLY Table 4.** Changes by Treatment Group in Women with Elevated Insomnia Symptoms at Screening

| | CBT+ | | | TAU+ | | | Group Difference | | |
|---|---|---|---|---|---|---|---|---|---|
| **Outcome Measure** | **Change** | **CI** | ***P*** | **Change** | **CI** | ***P*** | **Change** | **CI** | ***P*** |
| **Primary** | | | | | | | | | |
| Insomnia Severity Index | | | | | | | | | |
| Baseline to Post | -6.12 | -8.08, -4.15 | <.001 | -2.94 | -5.03, -0.86 | .006 | -3.17 | -5.95, -0.40 | .025 |
| Post to Follow-up | 0.54 | -2.04, 3.11 | .683 | -3.49 | -6.49, -0.50 | .022 | 4.03 | 0.09, 7.97 | .045 |
| **Secondary** | | | | | | | | | |
| Sleep-Related Impairment | | | | | | | | | |
| Baseline to Post | -5.98 | -8.74, -3.23 | <.001 | -2.38 | -5.36, 0.60 | .118 | -3.60 | -7.55, 0.34 | .073 |
| Post to Follow-up | -2.68 | -6.06, 0.71 | .121 | -2.69 | -6.58, 1.19 | .174 | 0.02 | -5.13, 5.16 | .995 |
| Sleep Disturbance | | | | | | | | | |
| Baseline to Post | -6.70 | -9.41, -3.98 | <.001 | -4.66 | -7.55, -1.78 | .002 | -2.03 | -5.84, 1.78 | .295 |
| Post to Follow-up | 0.90 | -2.65, 4.45 | .620 | -3.97 | -8.05, 0.10 | .056 | 4.87 | -0.52, 10.26 | .077 |
| Fatigue | | | | | | | | | |
| Baseline to Post | -6.31 | -8.90, -3.72 | <.001 | -0.91 | -3.73, 1.92 | .530 | -5.40 | -9.22, -1.59 | .005 |
| Post to Follow-up | -2.12 | -5.80, 1.56 | .258 | -1.45 | -5.63, 2.73 | .496 | -0.67 | -6.02, 4.68 | .806 |
| Depression Symptoms | | | | | | | | | |
| Baseline to Post | -0.55 | -3.14, 2.04 | .676 | -2.66 | -5.44, 0.12 | .061 | 2.11 | -11.60, 5.81 | .265 |
| Post to Follow-up | -2.64 | -5.56, 0.29 | .078 | -0.20 | -3.48, 3.09 | .906 | -2.44 | -6.87, 1.99 | .281 |
| Anxiety Symptoms | | | | | | | | | |
| Baseline to Post | -0.22 | -3.34, 2.90 | .890 | -2.70 | -6.10, 0.69 | .118 | 2.49 | -1.96, 6.93 | .273 |
| Post to Follow-up | -4.72 | -8.22, -1.21 | .008 | -1.01 | -5.12, 3.10 | .630 | -3.70 | -9.11, 1.71 | .180 |

*Note*. Results are in the subset of women who reported initial insomnia symptom scores of ≥ 8. CBT+ = cognitive behavioral therapy + bright light therapy. TAU+ = treatment as usual + relaxation audios.

**ONLINE ONLY Table 5.** Actigraphy and Sleep Diary Outcomes in Women with Elevated Insomnia Symptoms at Screening

| | Sleep Diary B (95% CI) | *P* | Actigraphy B (95% CI) | *P* |
|---|---|---|---|---|
| **Sleep Efficiency*** | | | | |
| *Intercept* | | | | |
| TAU+ | 41.09 (0.67, 2.24) | <.001 | 44.98 (40.91, 49.05) | <.01 |
| CBT+ | 47.09 (42.02, 52.16) | <.001 | 44.43 (40.66, 48.19) | <.001 |
| Group difference | 6.00 (-1.71, 13.71) | .127 | -0.55 (-6.10, 5.00) | .846 |
| *Slope* | | | | |
| TAU+ | 1.46 (0.67, 2.24) | <.001 | -0.14 (-0.94, 0.65) | .728 |
| CBT+ | 1.37 (0.58, 2.15) | .001 | 0.86 (0.07, 1.66) | .033 |
| Group difference | -0.09 (-1.20, 1.02) | .873 | 1.01 (-0.11, 2.12) | .078 |
| **Sleep Onset Latency*** | | | | |
| *Intercept* | | | | |
| TAU+ | 4.67 (3.86, 5.47) | <.001 | 1.53 (1.21, 1.86) | <.001 |
| CBT+ | 4.72 (4.02, 5.43) | <.001 | 1.44 (1.14, 1.74) | <.001 |
| Group difference | 0.06 (-1.01, 1.13) | .916 | -0.09 (-0.53, 0.35) | .682 |
| *Slope* | | | | |
| TAU+ | -0.10 (-0.22, 0.03) | .119 | -0.02 (-0.09, 0.05) | .594 |
| CBT+ | -0.13 (-0.25, -0.01) | .027 | -0.02 (-0.09, 0.05) | .512 |
| Group difference | -0.04 (-0.21, 0.13) | .679 | -0.00 (-0.10, 0.09) | .922 |
| **Wake after sleep onset*** | | | | |
| *Intercept* | | | | |
| TAU+ | 5.74 (4.63, 6.85) | <.001 | 6.94 (6.29, 7.59) | <.001 |
| CBT+ | 5.48 (4.51, 6.44) | <.001 | 7.26 (6.66, 7.86) | <.001 |
| Group difference | -0.26 (-1.73, 1.21) | .728 | 0.32 (-0.57, 1.21) | .482 |
| *Slope* | | | | |
| TAU+ | -0.12 (-0.25, 0.02) | .096 | 0.09 (-0.04, 0.21) | .168 |
| CBT+ | -0.20 (-0.34, -0.06) | .004 | -0.12 (-0.25, -0.00) | .046 |
| Group difference | -0.09 (-0.28, 0.11) | .380 | -0.21 (-0.38, -0.04) | .017 |
| **Total Sleep Time (hours)** | | | | |
| *Intercept* | | | | |
| TAU+ | 6.52 (6.07, 6.98) | <.001 | 6.96 (6.56, 7.35) | <.001 |
| CBT+ | 7.01 (6.61, 7.41) | <.001 | 7.38 (7.01, 7.74) | <.001 |
| Group difference | 0.48 (-0.13, 1.09) | .120 | 0.42 (-0.11, 0.96) | .122 |
| *Slope* | | | | |
| TAU+ | 0.16 (0.08, 0.23) | <.001 | 0.06 (-0.01, 0.12) | .083 |
| CBT+ | 0.06 (-0.02, 0.13) | .130 | -0.05 (-0.12, 0.01) | .113 |
| Group difference | -0.10 (-0.21, 0.01) | .065 | -0.11 (-0.20, -0.02) | .019 |

*Note*. Results are in the subset of women who reported initial insomnia symptom scores of ≥ 8.

*these measures were transformed, and results are on the transformed scales. CBT+ = cognitive behavioral therapy + bright light therapy. TAU+ = treatment as usual + relaxation audios.

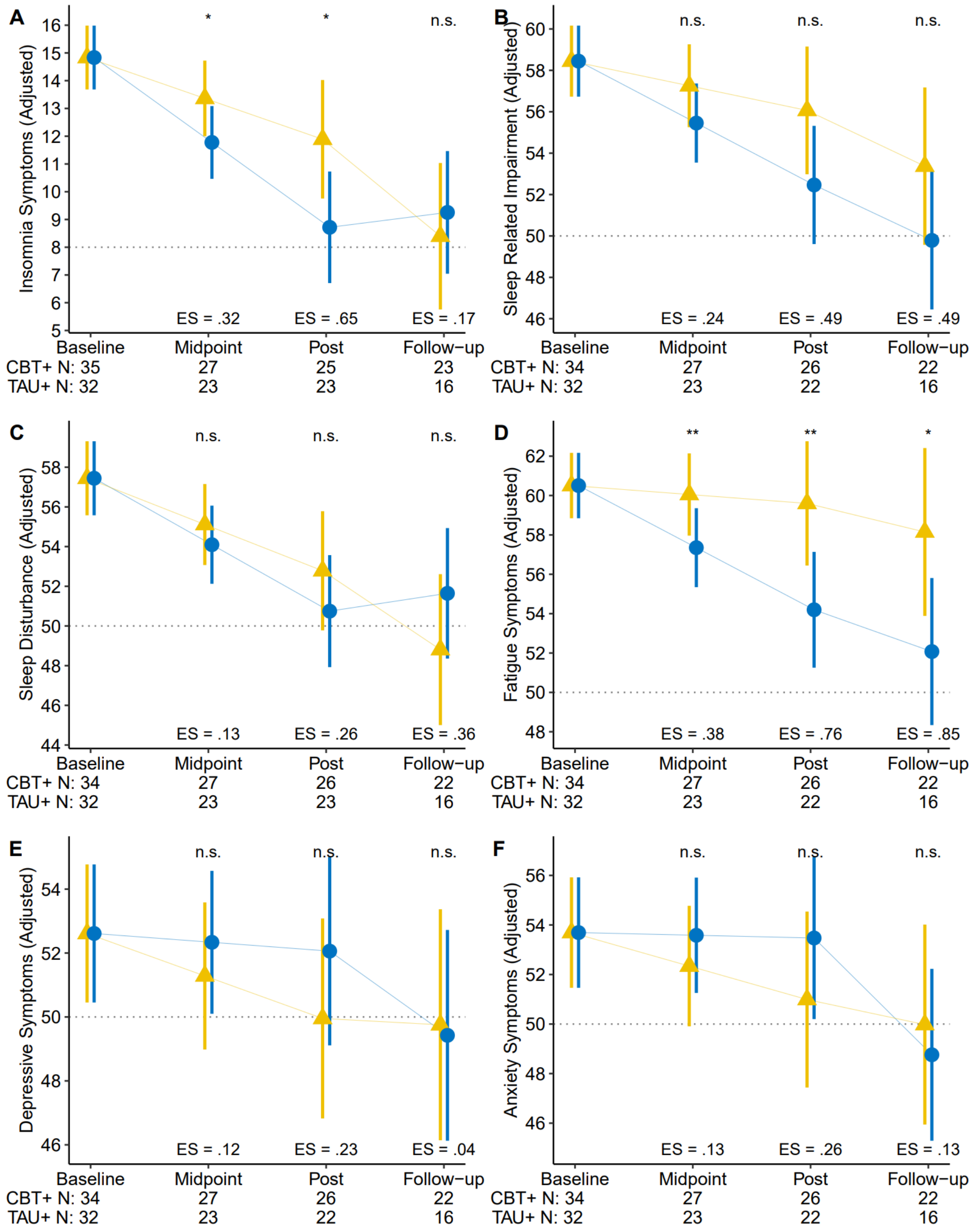

**ONLINE ONLY Figure 5**

Changes in mean patient reported outcome measures at baseline, mid-point, post-intervention and 3-month follow up by condition in those with initially elevated insomnia symptoms. Values are adjusted means and 95% confidence intervals from the primary analyses. Dashed grey line indicates population average values, where available, or minimal symptoms (insomnia symptoms only). The number of non-missing scores available in each group at each assessment are at the bottom. Baseline was completed just prior to week 0 (intervention start), midpoint at week 3, post at week 6, and follow-up 3-months after the completion of the intervention. CBT+ = cognitive behavioral therapy + bright light therapy. TAU+ = treatment as usual + relaxation audios. ES = effect size; *n.s.* = not significant; * $p < .05$; ** $p < .01$

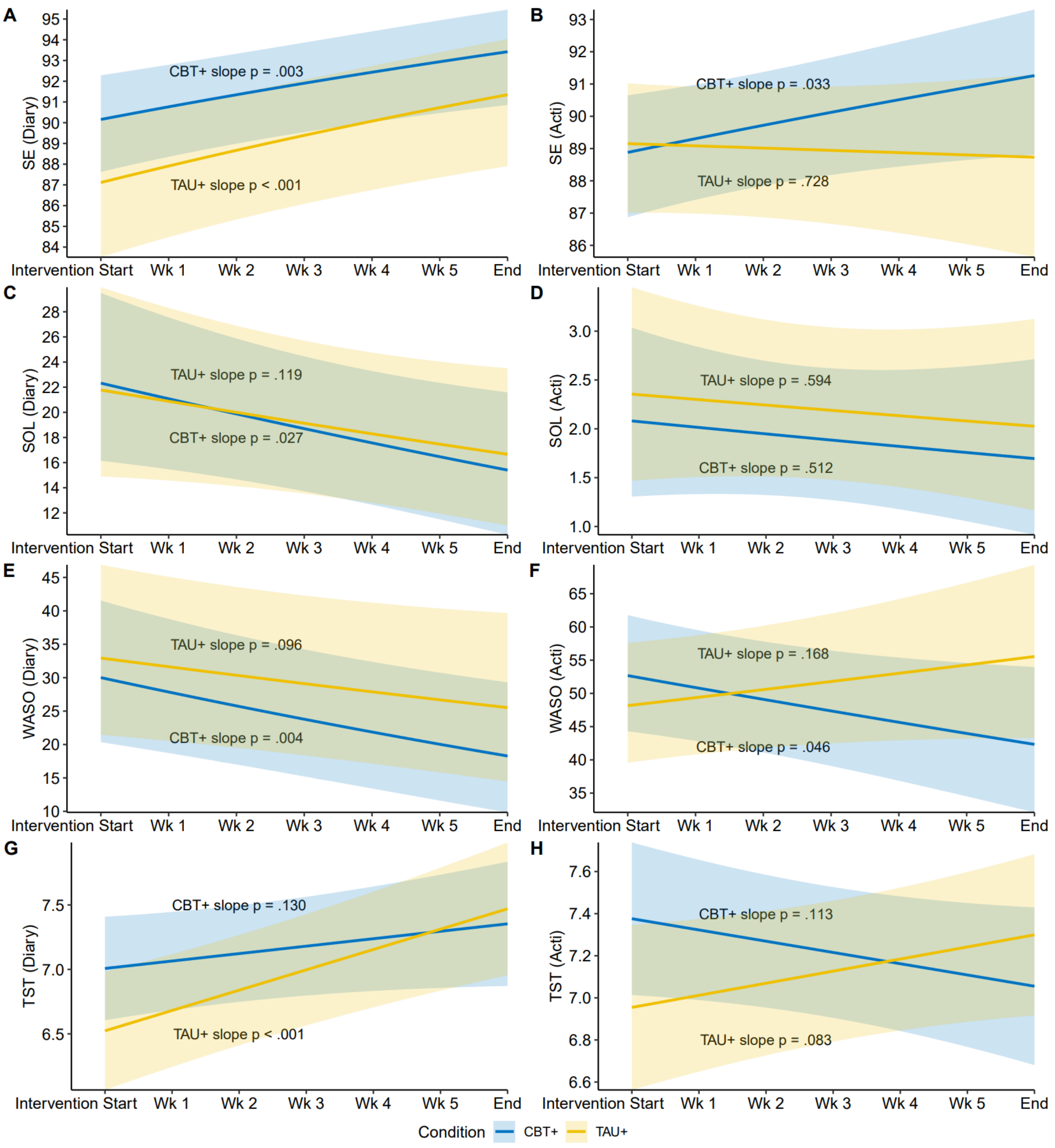

**ONLINE ONLY Figure 6**

Changes in objective and subjective sleep from start to end of intervention period by group. Results are in the subset of women who reported initial insomnia symptom scores of ≥ 8. Sleep diary and actigraphy assessments began the first night after the initial face-to-face intervention session. Graphs are estimated means over time with 95% confidence intervals shown in the shaded regions. Diary = sleep measures assessed by sleep diary. Acti = sleep measures assessed by wrist-worn actigraphy. SE = sleep efficiency, in percentages. WASO = wake after sleep onset, in minutes. SOL = sleep onset latency, in minutes. TST = total sleep time, in hours. CBT+ = cognitive behavioral therapy + bright light therapy. TAU+ = treatment as usual + relaxation audios.

**Exploratory Analyses – Factors Potentially Impacting Sleep**

Individual sleep patterns can differ on weekends [3], and insomnia is a known side effect of dexamethasone [4]. Daily dexamethasone use was reported by women in daily sleep diaries and the dates were logged. There were no differences between groups in the proportion of days of dexamethasone use reported nor in the number of weekend nights of sleep reported (Online Table 7).

In addition, at the immediate post intervention survey, women were asked to report on whether they used any other mental health or sleep treatment during the intervention. Frequencies and percentages for different mental health and sleep treatment are in Online Table 7. There were no differences in use of either treatments during the intervention by group.

Examining these variables in relation to sleep, on days when dexamethasone was used, women reported significantly lower $SE_{diary}$, longer $SOL_{diary}$, higher $WASO_{diary}$, and shorter $TST_{diary}$ (all $P < .001$). Similarly, on objective measures, $SE_{acti}$ was significantly lower ($P = .018$) and $TST_{acti}$ significantly shorter ($P < .001$) although there were no differences on $SOL_{acti}$ nor $WASO_{acti}$ (both $P > .10$).

On weekends versus weekdays, women reported significantly higher $SE_{diary}$ ($P < .001$), shorter $SOL_{diary}$ ($P = .028$), shorter $WASO_{diary}$ ($P = .005$), and longer $TST_{diary}$ ($P < .001$). However, on objective measures there were no weekend versus weekday differences on $SE_{acti}$, $SOL_{acti}$, nor $WASO_{acti}$ (all $P > .10$). $TST_{acti}$ was longer on weekends than weekdays ($P = .007$).

Given the robust differences in sleep on weekends versus weekdays and days when dexamethasone was taken, primary intervention efficacy analyses were conducted for all objective and subjective sleep parameters with weekend versus weekday nights and nights of dexamethasone use included as covariates in the analyses. Results from these analyses were largely similar to the main results from actigraphy and sleep diary data (Online Table 6). The patterns of change on each sleep parameter for both conditions (Online Figure 7), were similar to those shown in overall trial results. There were only slight changes. The improvement in $SE_{diary}$ in the CBT+ condition was no longer significant ($P = .13$) nor was the change in $TST_{diary}$ in the CBT+ condition ($P = 0.60$). The overall similarity between these results and the main sleep diary and actigraphy results indicates that weekend versus weekday and dexamethasone use did not meaningfully alter the effects of the intervention.

Finally, to explore whether receipt of mental health or sleep treatment during the intervention impacted outcomes, we examined whether our primary outcome, insomnia symptoms, differed at post intervention between women who did or did not receive different treatments using independent samples t-tests. There were no differences in insomnia symptoms at post intervention by receipt of psychological treatment for mental health (*P* = .25), attending groups or other supports (*P* = .41), medications for mental health (*P* = .72), psychological treatment for sleep (*P* = .63), or prescribed medication for sleep (*P* = .10). There was a significant difference between women who did not use over the counter or herbal sleep aids (M = 7.99) and those who did (M = 14.55) during the intervention (*P* = .001). However, women who reported using over the counter or herbal sleep aids during the intervention also differed on insomnia symptoms at baseline (*P* = .027). Further, the use of over the counter or herbal sleep aids was not associated with simple changes scores in insomnia symptoms from baseline to post intervention (*P* = .56). Therefore, we did not conduct further sensitive analyses for our primary efficacy analyses adjusting for other treatment receipt.

**ONLINE ONLY Table 6**. Actigraphy and Sleep Diary outcomes for both conditions, adjusting for weekday vs weekend and dexamethasone use.

| | **Sleep Diary B (95% CI)** | ***P*** | **Actigraphy B (95% CI)** | ***P*** |
|---|---|---|---|---|
| **Sleep Efficiency*** | | | | |
| *Intercept* | | | | |
| TAU+ | 44.30 (39.98, 48.61) | <.001 | 44.51 (41.22, 47.79) | <.001 |
| CBT+ | 52.10 (48.15, 56.04) | <.001 | 46.00 (42.79, 49.21) | <.001 |
| Group difference | 7.80 (1.96, 13.64) | .009 | 1.49 (-3.09, 6.08) | .52 |
| *Slope* | | | | |
| TAU+ | 1.43 (0.71, 2.15) | <.001 | -0.08 (-0.71, 0.54) | .80 |
| CBT+ | 0.57 (-0.16, 1.31) | .13 | 0.65 (-0.02, 1.32) | .06 |
| Group difference | -0.86 (-1.89, 0.17) | .10 | 0.12 (-0.19, 1.65) | .12 |
| **Sleep Onset Latency*** | | | | |
| *Intercept* | | | | |
| TAU+ | 4.39 (3.80, 4.99) | <.001 | 1.39 (1.15, 1.64) | <.001 |
| CBT+ | 4.18 (3.64, 4.73) | <.001 | 1.37 (1.12, 1.61) | <.001 |
| Group difference | -0.21 (-1.01, 0.60) | .61 | -0.03 (-0.37, 0.31) | .87 |
| *Slope* | | | | |
| TAU+ | -0.11 (-0.20, -0.01) | .03 | -0.01 (-0.06, 0.05) | .84 |
| CBT+ | -0.05 (-0.14, 0.05) | .33 | -0.04 (-0.10, 0.02) | .19 |
| Group difference | .06 (-0.07, 0.19) | .38 | -0.04 (-0.11, 0.04) | .40 |
| **Wake after sleep onset*** | | | | |
| *Intercept* | | | | |
| TAU+ | 5.37 (4.56, 6.18) | <.001 | 7.10 (6.57, 7.62) | <.001 |
| CBT+ | 4.65 (3.92, 5.39) | <.001 | 7.04 (6.52, 7.55) | <.001 |
| Group difference | -0.72 (-1.81, 0.37) | .20 | -0.06 (-0.79, 0.68) | .88 |
| *Slope* | | | | |
| TAU+ | -0.15 (-0.27, -0.03) | .01 | .04 (-0.05, 0.14) | .37 |
| CBT+ | -0.09 (-0.21, 0.03) | .16 | -0.09 (-0.20, 0.01) | .08 |
| Group difference | 0.06 (-0.11, 0.24) | .45 | -0.14 (-0.28, 0.01) | .06 |
| **Total Sleep Time** | | | | |
| *Intercept* | | | | |
| TAU+ | 6.68 (6.31, 7.04) | <.001 | 7.00 (6.66, 7.34) | <.001 |
| CBT+ | 7.36 (7.03, 7.69) | <.001 | 7.42 (7.09, 7.75) | <.001 |
| Group difference | .68 (0.19, 1.17) | .01 | 0.42 (-0.05, 0.89) | .08 |
| *Slope* | | | | |
| TAU+ | 0.13 (0.06, 0.19) | <.001 | 0.01 (-0.05, 0.08) | .63 |
| CBT+ | 0.02 (-0.05, 0.08) | .60 | -0.05 (-0.12, 0.01) | .13 |
| Group difference | -0.11 (-0.20, -0.02) | .02 | -0.07, (-0.15, 0.02) | .15 |

*Note*. *these measures were transformed, and results are on the transformed scales. CBT+ = cognitive behavioral therapy + bright light therapy. TAU+ = treatment as usual + relaxation audios.

**ONLINE ONLY Table 7.** Variables Potentially Impacting Sleep During the Intervention by Group

| | CBT+ | TAU+ | *P* | ES |
|---|---|---|---|---|
| Days of Dexamethasone Use, *N (%)* | 176 (11.7) | 126 (9.3) | .20 | – |
| Weekend Nights of Sleep, *N (%)* | 428 (28.5) | 386 (28.3) | .90 | – |
| Mental Health Treatment (Yes), *N (%)* | | | | |
| Psychological (Yes), *N (%)* | 11 (30.6) | 6 (18.8) | .26 | .14 |
| Group or Other Support Resource (Yes), *N (%)* | 9 (25.0) | 4 (12.5) | .19 | .16 |
| Medication (Yes), *N (%)* | 5 (13.9) | 3 (9.4) | .56 | .07 |
| Any Sleep Treatment (Yes), *N (%)* | | | | |
| Psychological Sleep Treatment (Yes), *N (%)* | 3 (8.3) | 1 (3.1) | .36 | .11 |
| OTC/Herbal Sleep Aid (Yes), *N (%)* | 6 (16.7) | 5 (15.6) | .91 | .01 |
| Sleep Medication (Yes), *N (%)* | 9 (25.0) | 8 (25.0) | >.99 | <.01 |

*Note*. P-values come from mixed effects logistic regression to account for repeated measures for days of dexamethasone use and weekend nights of sleep and chi-square tests for mental health and sleep treatment variables. ES = effect sizes, which are Cramer's V for categorical variables and are not available for repeated measures data. OTC = over the counter; N = 68 for post intervention. CBT+ = cognitive behavioral therapy + bright light therapy. TAU+ = treatment as usual + relaxation audios.

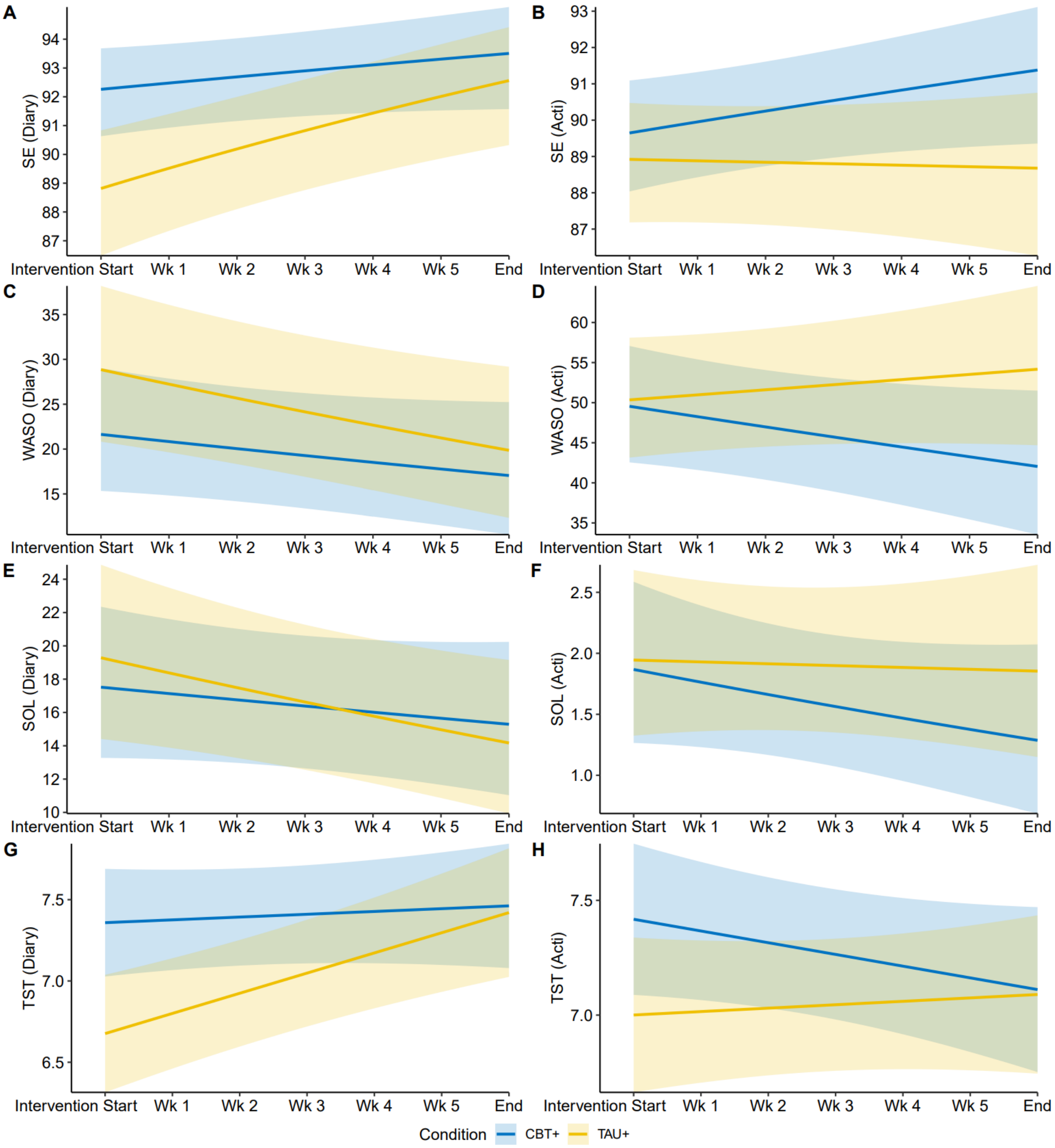

**ONLINE ONLY Figure 7**

Changes in objective and subjective sleep from start to end of intervention period by group with weekend versus weekday and daily dexamethasone use included as covariates. Sleep diary and actigraphy assessments began the first night after the initial face-to-face intervention session. Graphs are estimated means over time with 95% confidence intervals shown in the shaded regions. Diary = sleep measures assessed by sleep diary. Acti = sleep measures assessed by wrist-worn actigraphy. SE = sleep efficiency, in percentages. WASO = wake after sleep onset, in minutes. SOL = sleep onset latency, in minutes. TST = total sleep time, in hours. CBT+ = cognitive behavioral therapy + bright light therapy. TAU+ = treatment as usual + relaxation audios.
